# Community Engagement in Dengue Interventions in Conflict-affected Aden, Yemen: An Implementation Research Using CFIR Framework 2.0.

**DOI:** 10.1101/2024.12.30.24319744

**Authors:** Mustafa Mohammed Dhaiban, Huda Basaleem, Neetu Purohit

**Affiliations:** Indian Institute of Health Management Research University: IIHMR University; Community Medicine and Public Health Department, Faculty of Medicine and Health Sciences, University of Aden, Aden, Yemen

## Abstract

Dengue fever is described by the World Health Organization (WHO) as “the most important mosquito-borne viral disease in the world.” In Yemen, dengue cases have consistently increased, with Aden Governorate reporting 12% of total cases and over 45% of dengue-related deaths, making it a leading public health challenge. Conflict has exacerbated health system weaknesses, limiting resources and energy for prevention efforts. Community engagement (CE) emerges as a critical approach to address these challenges.

**This study aimed** to identify barriers and facilitators to CE in dengue interventions within conflict-affected contexts and recommend evidence-based strategies for improved implementation. A cross-sectional qualitative study was conducted between February and July 2024 in Aden Governorate, involving 15 in-depth interviews (IDIs) and four focus group discussions (FGDs) with key stakeholders.

**Findings** revealed that while stakeholders perceived CE interventions positively, challenges included systemic issues such as lack of community trust, resource shortages, and health system politicization. Conflict intensified these barriers, reflected in security concerns, inadequate training, and fragmented institutional work. Financial constraints also limited implementers’ capacities. Facilitators included moral incentives, personal recognition, and community support from business owners. However, the dominance of policymakers and influencers due to systemic weaknesses negatively impacted the implementation process, particularly in participant selection and coordination strategies.

**In conclusion,** addressing systemic fragmentation, enhancing political support, and defining clear coordination plans are critical to adapting CE interventions effectively in conflict

**Author Summary:** Dengue fever is considered a major health problem for many low-and middle-income countries, leading to high numbers of mortality and morbidity rates with a significant economic burden. Our study highlights the intersection of public health challenges and conflict dynamics, providing unique insights into how community engagement strategies for vector-borne diseases can be adapted to conflict settings. Using the implementation research tool CFIR Framework 2.0, we identified critical barriers, including systemic fragmentation, lack of trust, resource shortages, and weak coordination mechanisms. Conversely, facilitators such as moral incentives, personal recognition, and community-driven support offer actionable strategies for improving implementation outcomes.

## Introduction

Dengue Fever is a global health concern affecting over 100 countries, causing approximately 100 million cases and 25,000 deaths annually (1), the WHO describes dengue as “the most important *mosquito-borne viral* disease in the world”(2). In southern Yemen, the population has been severely impacted by dengue fever, with 104,940 reported cases and 221 deaths between 2020 and August 7, 2024. In Aden governorate alone, over 12,203 cases and 100 deaths have been recorded during this period.(3). Yemen suffers greatly from dengue, especially because of the ongoing political conflict and instability that have weakened its healthcare system and exacerbated the multifaceted nature of dengue disease (4–8).

Publication highlighted the connection between the presence of Internally Displaced Persons (IDPs) and increased dengue incidence (8). A systematic review by *Mahmud and others* (9) emphasize the effectiveness of combining environmental management and community participation in dengue control. Moreover, intersectoral coordination and support from various agencies have shown success in sustaining environmental management (6,10). Numerous national research studied knowledge practice and attitude (KAP) shows clear gap between the community knowledge and their commitment to preventive measures (11–15). Study conducted in AL-Hodeidah governorate, shows 57% from the participants believed that eliminating dengue breeding sites is government responsibility (12).However, research highlighted the importance of CE as a key solution to health issues within the current Yemen situation to assure the continuity of the health message (14,16).

An unpublished report for the Mentor Initiative an intervention implemented across five districts in Aden Governorate. The project included various activities, including door-to-door education and entomological surveys and Information Education and Communication (IEC) mass media campaigns. The main challenges reported included delays in coordination due to COVID-19, the majority of government employees taking their annual leaves during Ramdan, high temperature and humidity making it difficult to finding *mosquito pupae*, difficulties in training health volunteers, pressure from beneficiary for material support, challenges in obtaining epidemiological data from responsible authorities and delay of getting permeation for the IEC approvals (17).

The objective of this study was: to determine the barriers and facilitators for CE in dengue interventions and recommend evidence-based strategy for better implementation.

## Methods

This is cross-sectional study using qualitative approach, conducted from February 2024 to July 2024. The study targeted key stakeholders involved in CE activities related to dengue prevention and control. Purposive sampling with snowball technique was used to identify stakeholders (18). The qualitative component included 15 (IDIs) and 4 (FGDs) to reach the saturation point.

### Study Setting

Aden governorate is the largest city of southern Yemen and it is also the administrative center, since 2015, It is positioned along the coast of the Gulf of Aden, covering a total area of 712.48 km². This city falls within the tropical region (19). Aden is temporarily internationally recognized capital of Yemen, with a population of approximately 1.14 million residents (20).

### Implementation framework

The CFIR Framework is one of the commonly used tools in implementation science, which consist of five domains: The Innovation Domain examines key stakeholders’ perceptions of the CE intervention. The Outer Setting Domain identifies macro-level factors that influence the intervention and shape the inner setting. The Inner Setting Domain focuses on the micro-level environment where the CE intervention is implemented. The Individual Domain considers the roles and characteristics of individuals involved in the intervention. Finally, the Implementation Process Domain captures the processes associated with implementation, (21). The use of CFIR framework started by selecting the constructs and sub-construct that are related to the research objective which where 23 construct out of the total 48, and 5 sub-construct out of the total 19 (22,23). In this implementation research, the CFIR framework was utilized to identify barriers and facilitators associated with the current design of CE interventions for dengue control, which primarily relies on direct communication with community through door-to-door education by health workers and community volunteers

### Data collection

The qualitative data for (IDI) (Annex 1) and (FGD) (Annex 2) were collaboratively conducted by the principal investigator and assistant possessing a good background in qualitative data methodology. The process guided by CFIR 2.0 framework interview guideline tool, initially developed from CFIR.com (22). The interview protocol further refined to integrate specific questions from the CFIR 2.0 framework that directly pertain to the research questions and objectives. Table No.1 present the CFIR framework domains and construct, along with the distribution across each IDI and FGD.

**Table No. (1):** Distribution of CFIR Constructs Across IDI and FGD.

| **CFIR Domain** | **Construct** | **Included in IDI** | **Included in FGD** |
| --- | --- | --- | --- |
| Innovation Domain | Evidence-Base | Yes | Yes |
|  | Relative Advantage | Yes | Yes |
|  | Innovation Complexity | Yes | Yes |
|  | Innovation Cost | Yes | No |
| Outer Setting Domain | Critical Incidents | Yes | Yes |
|  | Local Attitudes | Yes | Yes |
|  | Local Conditions | Yes | Yes |
|  | Partnerships & Connections | Yes | Yes |
|  | Policies & Laws | Yes | No |
|  | Financing | Yes | No |
| Inner Setting Domain | Work Infrastructure | Yes | No |
|  | Relative Priority | Yes | No |
|  | Incentive Systems | Yes | Yes |
|  | Available Resources | Yes | Yes |
|  | Access to Knowledge & Information | Yes | Yes |
| Individual Domain | Policymaker need, capability, motivation | Yes | Yes |
|  | Implementation team Capability, motivation | Yes | Yes |
|  | Opinion Leaders | Yes | Yes |
| Implementation processes domain | Teaming | Yes | Yes |
|  | Assessing Needs | Yes | No |
|  | Planning | Yes | No |
|  | Tailoring Strategies | Yes | No |
|  | Adapting | Yes | Yes |

### Data analysis

The qualitative guides were developed in English and translated into Arabic for interviews. After obtaining informed consent, interviews were recorded, and then coded using NVivo 14 software. Content deductive analysis, by CFIR 2.0 codebook guided the identification and analysis of key themes relevant to the research objective (24,25).

## Results

### Participants description

The participants category and their number and sex, roles and years of experience, are detailed in table No. (2).

**Table No. (2).** Participants profile.

| **Participant Category** | **Participant No. (Sex)** | **Participant role (years of experience)** |
| --- | --- | --- |
| **Organization consultant category** | **P 7** (Male) | Public Health and Epidemiology Consultant (35 years) |
|  | **P 15** (Male) | Public Health Consultant on SBC (30 years) |
|  | **P 5** (Male) | Public Health Consultant on vector-borne disease (5 years) |
| **Organization employees** | **P 6** (Male) | International Organization Employee on vector-borne disease (2 years) |
|  | **P 12** (Female) | Executive Director of local organization worked at Aden vector-borne disease (14 years) |
| **Community Committee** | **FGD 1** (Female) | Male and Female Participants are part of community committee and involved at different roles such as women and chilled officers, secretaries, mediator and resolving family dispute and conflict at Dar Sa'ad district |
|  | **FGD 2** (Male) |  |
|  | **FGD 3** (Female) | Male and Female Participants are part of community committee and involved at different other roles such as leadership and governess, support community initiative, health campaign. at AL-Mansoura district |
|  | **FGD 4** (Male) |  |
| **Ministry of public Health and population (MoPHP) Employee** | **P 4** (Female) | Employee at ministry level on epidemiological surveillance (3 years) |
|  | **P 8** (Male) | Employee at ministry level on national program for health education and information. (10 years) |
|  | **P 9** (Male) | Employee at ministry level on national program for health education and information (21 years) |
|  | **P 10** (Female) | Employee at Aden health office - Governorate level (15 years) |
|  | **P 11** (Male) | Employee at Aden health office - Governorate level (11 years) |
|  | **P 13** (Male) | Employee at Aden health office – AL-Mansoura district (6 months) |
|  | **P 14 (Male)** | Employee at Aden health office Al-Mansoura District Dar Sa'ad district (2 years) |
|  | **P 1 (Male)** | National Malaria Control Program Employee (17 years) |
|  | **P 2 (Male)** | National Malaria Control Program Employee (25 years) |
|  | **P 3 (Male)** | National Malaria Control Program Employee (1 year) |

#### 1. Innovation Domain

##### This theme is to investigating perceptions of key stakeholders about the component of community engagement intervention

All participant categories emphasized the importance of CE for dengue prevention, viewing it as a key social and behavioural intervention. A participant from the National Malaria Control Program (NMCP) with over 17 years of experience stated: *“Awareness-raising and community involvement in dengue control are crucial because we cannot fully control the vector or guarantee complete treatment.”* Additionally, participants shared personal experiences from previous health interventions. Participant No. 12 described an innovative approach in Lahj Governorate: *“We involved elementary schools by asking fifty students to collect water samples from their homes to identify places where water accumulates and bring samples of larvae.”*

However, most participant categories identified trust as a significant challenge, with the exception of those from the organization employee category. Organization consultants and community committee members pointed to rumors stemming from vaccine campaigns as a source of distrust. Community committee participants further connected trust issues to the quality of health services, including doctor-patient relationships, drug pricing, and hygiene standards at health centers. A male community committee member from Al-Mansoura explained: *“The community has lost trust due to the perceived relationship between doctors, pharmaceutical companies, and financial benefits, leading to a lack of confidence.”* Another participant added: *“We found a pile of discarded medicines in the garbage. Upon investigation, we discovered they belonged to a company with offices outside the governorate.”*

Most MoPHP participants identified financial constraints and rumors as the primary challenges to CE efforts. Meanwhile, organization participants regarded CE interventions as low-cost initiatives, with the primary expenses involving brochures, transportation, and volunteer incentives. Participant No. 15 reflected: *“Involving the community is both easy and difficult at the same time, but it is considered the cheapest option.”*

#### 2. Outer setting Domain

##### This theme is to determine the macro-level elements that influence intervention and shaped of or from inner setting theme

All participant categories noted that the political situation has led to insecurity, and climate change has caused rainfall and flooding, which they consider critical incidents. Additionally, all categories except organization employees viewed rumors and mistrust stemming from social media and vaccine-related misinformation as significant issues. However, organization employees and community committee members focused on the duality of authority and the challenges in obtaining required permits as critical incidents a participant No. 12 stated: *"Armed conflicts are considered the most important obstacles, along with the accompanying duality of authority. Each authority considers the activity to be its own, whether it is water management, agriculture."*

All participant categories agreed on the positive attitude of the community in Aden and their willingness to engage, noting that economic constraints affect the extent of their engagement. The low economic situation was seen as a major factor limiting CE. Participants explained that financial hardship prioritizes other needs over health and influences decisions around seeking treatment and following preventive measures male community committee said*:” The impact is undoubtedly negative. Many people have limited incomes and cannot afford medication. They resort to herbal remedies, which affects their health. I had to get it myself”*. Community committee members also highlighted shifts in community behavior since the war, with an increased tendency to request financial or material assistance. Participants debated the root causes of this change created by the conflict or negative humanitarian interventions design. Participant No.11 said: “*In comparison to before 2015, awareness focused on natural satisfaction, environmental health, extracting birth certificates, education, school dropouts, healthy nutrition, and everything that builds human health and development. There were no great material demands like now due to displacement, aid, and poverty.*”. Similarly, participants No.2 and No.3 from community committee of AL-Mansoura district have said: “*After the war, people became more materialistic. As community committees, our work is voluntary, and people constantly inquire about compensation”* and after he finished the participant No.3 responded to his point *by “I don’t believe people have become inherently materialistic. The situation forced them to be this way. The situation was good before the war. After the war, prices skyrocketed.”* Finally, participant No.5 working as organization consultant believes the community behaviour changed due to relief interventions from organization aid. *“We also found that the community’s response to many awareness projects after 2015 was affected by other relief interventions, and their demands for in-kind assistance and other things became greater than their interest in health awareness”*

Participants agreed on the impact of the political situation, though specific concerns varied. Organization consultants highlighted the deviation in health policies between the internationally recognized government and the de facto Houthi authorities, while organization employees focused on the influence of political parties seeking benefits from interventions. Community committee members supported these points, mentioning how political and regional backgrounds influence health system hiring practices. MoPHP participants echoed these concerns, with Participant No. 1 noting, *“The more politically frustrating the general situation is, the more it reflects on the community’s psyche and their receptiveness to dengue prevention education.”* Additionally, a members of community committee said : *“Politicization of healthcare is a problem. It becomes a battleground for different parties. For instance, if a particular party controls the health sector, individuals manipulate it for their party’s benefit. This is wrong”*

The organization category emphasized the importance of international organizations working with multiple government and local partners, typically through health clusters. Although partnerships with local organizations are generally positive, issues with collaboration and trust between government bodies persist. Participant No. 15 stated, *“We still need the government side to feel mutual trust because the absence of trust within the institutional system is a problem. When they say they cannot transfer millions to the district account due to monitoring and audit limitations, and prefer transferring it to the governorate to oversee as a partner, it shows a lack of trust.”* Community committee opinions on partnerships with health authorities varied across districts, while MoPHP participants acknowledged positive partnerships with organizations but pointed to internal connection issues. However, with problems of connection and partnership a new initiative shows some improvement. The initiative conducted by the national program for health education and information using WhatsApp groups to communicate with different stakeholders.

All participant categories indicated a lack of awareness of policies and laws related to CE. Organization consultants referenced old regulations from 30 years ago, which defined the CE process in hospitals, as well as the public health law, which includes community involvement. Currently there are general custom regulate community volunteer work and CE in general. With new regulation from Aden governor to establish community committee work as partnership between community and Aden governorate. And the participant No.13 working in the health office of Al-Mansoura District said*: “As for us, the establishment of community committees by a decision from the governor of the governorate. They have a central department in the governorate and encourage their participation in any health aspect.”*

Most participants considered international aid the primary funding source for CE interventions. Nevertheless, Interrupted funding from donors due to changing priorities or emergency situations was another concern. Participant No 5 stated: “*Sudden cessation of support due to weak funding or changing priorities of implementing organizations, such as what happened during the Corona pandemic, with some funding for specific activities being diverted to activities related to COVID-19.”* Organization participants emphasized the need for evidence-based data and high-quality proposals, while the government has not prioritized CE interventions. However, MoPHP participants (Nos. 1, 2, and 13) suggested that business owners and charities could potentially serve as alternative funding sources the participant No.13 said: *“Community participation interventions can be implemented through charities, businessmen, and financiers. As for me and my work in the health centre, there was sympathy from businessmen and financiers. During Ramadan, I was able to set up a Ramadan medical camp for people with chronic diseases and dispensed free medicines at a cost of 20 million Yemeni rials.”*.

#### 3. Inner Setting Domain

##### The inner setting referring to the micro-level where the CE intervention implemented

In the inner setting domain, all participant categories agreed on the structural weaknesses within government sectors and noted conflicts of interest between the Ministry of Public Health and Population (MoPHP) and the Aden health office, as each attempt to leverage institutional weaknesses for its own advantage. Participant No. 11 observed: *“There was previously confusion about whether activities in Aden could be carried out by the ministry because it was new. However, the vision became clear that the governorate has specific tasks and geographical boundaries.”* Additionally, differences in selection criteria between districts, favouritism, financial motives, and interference from influential figures were noted as challenges in workforce selection.

All participants recognized the priority of CE interventions and the increasing interest from international organizations in SBC interventions. However, there were differences in priorities among MoPHP national programs. As Participant No. 8 noted, *“In reality, the National Malaria Control Program focuses more on the control aspect, with awareness seen as secondary unless there is an increase in funding specifically for it. Otherwise, they turn to fogging."* This emphasis on control affects the work of funding organizations; for example, WHO supports prevention mainly through fogging and bed net distribution, whereas UNICEF prioritizes CE and SBC interventions for diseases beyond vector-borne illnesses. Participant No. 1 from NMCP commented: *“Community engagement is a priority, but donor agencies are slow to fund it even when outbreaks occur. By the time we try to involve the community, the issue has already diminished due to climate factors, and it becomes difficult for community actors to effectively communicate the message."*

Participants agreed on the effectiveness of intangible incentives, such as training workshops and certificates of appreciation, for motivating healthcare and community workers. CE interventions for dengue control, however, rely heavily on community volunteers, who are paid for their efforts. Participant No. 15, an organization consultant for SBC, expressed concerns that this practice undermines the principle of volunteering and can lead to financial burdens: *“We don’t just need to increase incentives; we need to rethink volunteer work. For instance, if imams or teachers who already receive a salary take responsibility for community awareness, we could reduce volunteer costs.”*

The Community Committee and MoPHP participants noted that the low economic situation necessitates financial incentives for volunteers. All participant categories agreed on the lack of training and guidelines specific to CE and SBC interventions, and most emphasized the role of business owners and charity organizations, though opinions within MoPHP varied. While some participants (Nos. 4, 8, 9, and 3) believed there were limited resources, others (Nos. 10, 13, 14, 1, and 2) saw opportunities in charity, business sponsorship, and the use of social and government media to support these interventions.

#### 4. Individual domain

##### This theme is focusing in the roles of individuals and characteristic of people involved at CE interventions by understand their needs, capabilities, and motivations. It divided into three main categories policymakers, implementers, and influencing or opinion leaders

All participant categories believed that CE interventions aligned with policymakers’ needs and served their interests, as they could help reduce dengue morbidity and mortality. However, Participant No. 15, an organization consultant, expressed doubts about the implementation aspect due to the financial burden, stating, *“The decision for community engagement interventions serves them, but unfortunately, the implementation doesn’t. Each of them might see this as a burden on their ministry, despite knowing they need each other.”* Participants from the MoPHP category viewed the decision to implement CE as a multisectoral responsibility. Conversely, the community committee category believed that it was primarily the responsibility of MoPHP, followed by the Aden health office and local authorities, as Participant No. 4 explained: *“The local authority, the Public Health and Population Office, the Health Office at the district level, and the community committees are responsible for community engagement interventions.”* Consequently, participants identified health volunteers and community committees as key players in implementation, emphasizing the importance of collaboration with the governorate, district health offices, and related sectors. They also recognized influential individuals, including the Prime Minister’s office, Aden’s local council, and the governor.

Organization consultants perceived a lack of capability among policymakers, as Participant No. 7 stated: *“I think there is a gap here, particularly regarding the understanding of new approaches like behavior change communication.”* Conversely, other participants had a positive view of policymakers’ capabilities, with some commenting, *“Certainly, they wouldn’t have reached these positions without the necessary skills and knowledge.”* Opinions varied on the capabilities of implementers as well. While some believed them to be capable, others felt they needed additional training. Participant No. 12, an executive director for a local organization, remarked, *“Government officials are experienced in their fields. In organizations, the situation varies; some may be experienced and competent, while others may be young and hardworking. But as government officials, they don’t reach positions like sector managers or center heads without experience.”*

Views on policymakers’ motivation were mixed. Some felt that policymakers lacked motivation, while others believed they were motivated by financial incentives or by epidemiological needs. A female community committee member commented, *“They are only motivated when external entities like NGOs incentivize them.”* On the other hand, participants generally saw implementation teams as motivated, with Participant No. 7, an organization consultant, noting the impact of financial incentives and epidemiological conditions: *“When individuals and families experience pain through these infections, their level of enthusiasm somewhat increases. However, their motivation and readiness surge even more when their needs are prioritized. Additionally, their personal income rises when they work in the field.”*

Participants also felt that religious leaders held influence over CE efforts. Participant No. 1, who has worked at the NMCP for 20 years, emphasized the role of financial factors as a major influence, stating, *“The influencers are stakeholders with financial interests. They will strongly encourage and support the work, but if there are no benefits, they may push for the complete cancellation of the implementation. This applies to the entire health pyramid, from the ministry to the National Malaria Control Program, down to the base.”*

#### 5. Implementation Process Domain

##### captures the processes associated with implementation

All participant categories identified interference in participant selection as a major challenge affecting team dynamics during implementation. Participant No. 2 remarked: *“What is happening now in terms of interference in the selection of candidates by influential parties directly reflects negatively on the quality of work and the effectiveness of delivering the message.”* In addition, participants agreed that preparedness before emergencies and coordination were weak. As Participant No. 10 stated: *“Cooperation, partnership, and coordination occur when the alarm bell rings. Efforts are mobilized, led by the governor, and an operations room is formed with each party doing its work only when there is danger.”* All participants agreed on the need for a well-defined plan with specific objectives and indicators for CE in dengue interventions. Participant No. 6 emphasized this, stating*: “The top priority is to set specific goals and outcomes to monitor with accurate indicators, helping us analyze implementation correctly.”* MoPHP employees also highlighted the importance of proper training in CE engagement and SBC concepts, as well as effective coordination policies between governmental and non-governmental sectors. Some MoPHP participants noted issues with unrealistic planning that is often not implemented, while others pointed to internal management challenges. Participant No. 8 shared, *“We prepared a plan with a national expert a year ago, but due to internal administrative disagreements, the workshop to discuss this strategy was delayed.”*

All participant categories stressed the importance of a strategic coordination plan participant No. 1 highlighted the value of collaboration across health programs: “*Activating the education and awareness department within the National Malaria Control Program, maintaining continuous media communication, and working with implementers in immunization and nutrition programs to deliver short awareness messages during their activities.”*. The MoPHP category further emphasized the need for political support and suggested leveraging available human resources, such as community committees, mosque leaders, students, teachers, and private sector funding. For adapting CE interventions, all participant categories agreed on the importance of ensuring funding availability. Organization participants believed this could be achieved with community ownership of CE interventions. Meanwhile, the Community Committee and MoPHP categories supported the need for dedicated funding allocations and suggested involving students, mosque leaders, and media channels including television, newspapers, radio, and social media also important of mentoring and accountable.

The tables below provide key insights: Table 3 summarizes the interrelations between the Implementation Process domain and other CFIR domains, while Table 4 highlights the main barriers and facilitators identified across CFIR domains.

**Table No (3):**
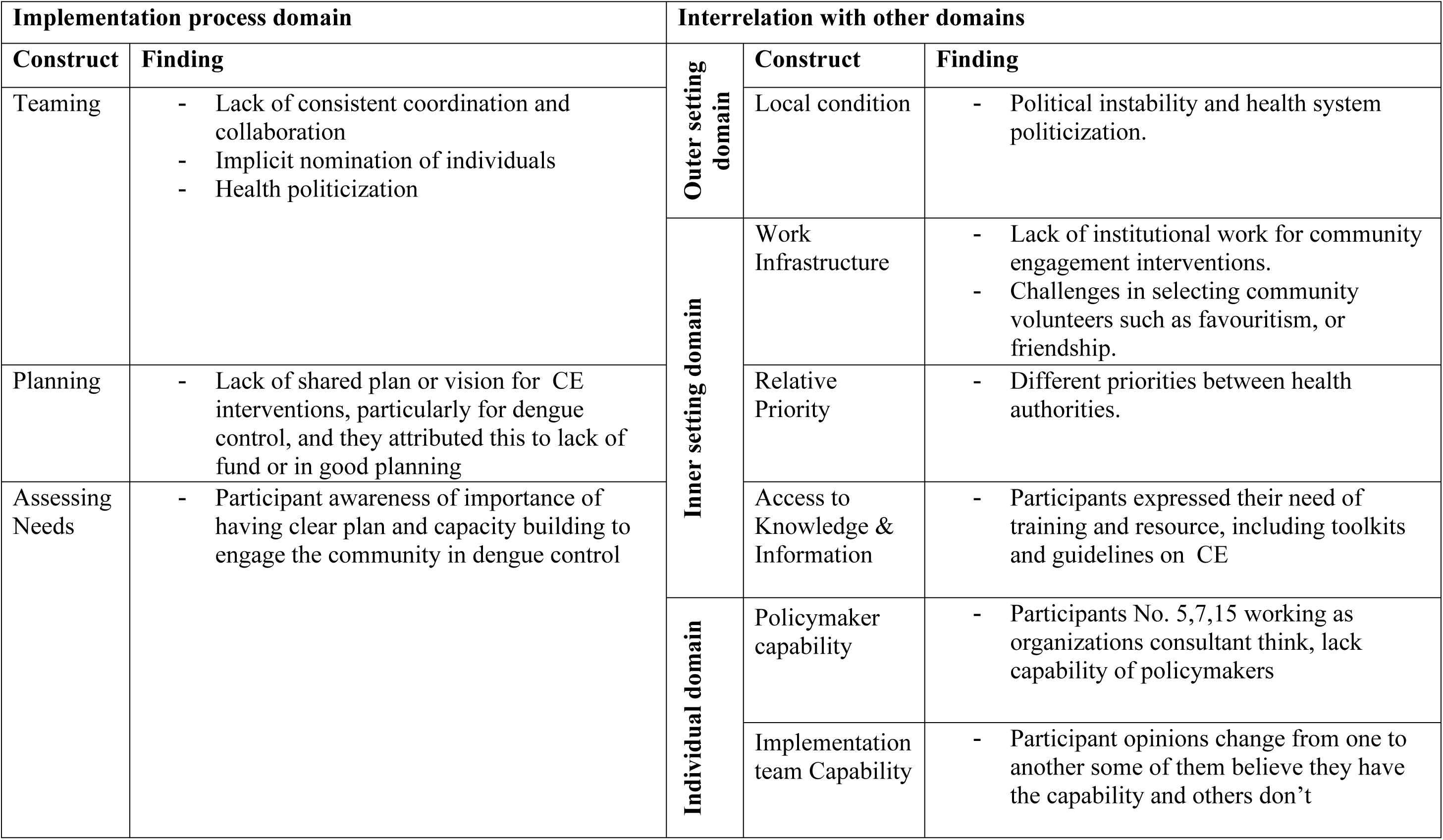
Interrelations Between Implementation Process Constructs and Other CFIR Domains.

**Table No (4):** Barriers and Facilitators Identified by CFIR Domains.

| **CFIR Domain** | **Barriers** | **Facilitators** |
| --- | --- | --- |
| **Innovation Domain** | • Distrust comes from rumours and low health services | • Agreement about the important of CE interventions<br>• Good personal experiences<br>• There is no complexity at CE interventions<br>• low-cost intervention |
| **Outer Setting Domain** | • Insecurity<br>• Climate change<br>• Duality of authority<br>• Bad economic status.<br>• Negative behaviour changes for community response<br>• Political instability and health system politicization.<br>• Inactivated legalization policy related to CE<br>• Interrupted funding from donors and changing priorities<br>• Lack of interest and vision for CE from MoPHP<br>• Low quality of interventions proposals. | • Collaborative attitude of the community<br>• Health clusters<br>• Partnerships with organizations are generally positive<br>• New technological initiative ( WhatsApp)<br>• New regulation of community committee from Aden governor.<br>• Funding from the international organization.<br>• Success experiences of business owners and charity |
| **Inner Setting Domaine** | • Lack of institutional work for CE interventions<br>• Issues in workforce selection.<br>• Difference in priorities between health authorities<br>• Depending on community volunteers.<br>• Depending on international fund<br>• Lack training and toolkits and guidelines for community engagement | • Good understanding to the importance of community engagement<br>• Moral incentives and personal recognition<br>• Funding resources through taxes, and f business owners and charity.<br>• Participant awareness of training gab. |
| **Individual Domain** | • Implementation aspect doesn't align with policymakers because of financial burden<br>• Lack of CE capabilities among policymaker (Organization consultants prospective)<br>• Fluctuated opinion about policymaker motivation<br>• Fluctuated opinion about Implementation team Capability | • CE intervention aligned with policymakers needs<br>• Good impression about policymaker capabilities (Community prospective)<br>• Implementation team are generally motivated with effect of the financial return, epidemiological situation<br>• Prime minister office and Aden governorate local council, religious people have the current influence. |

## Discussion

### Innovation domain

While there are national evidence supports CE (the Innovation) (27) and international evidence highlights the effectiveness of CE interventions for dengue control (28), the available evidence emphasizes the importance of CE in conflict and post conflict context (29,30) specifically, it underscores the significance of CE approaches in preventing infectious disease and dengue epidemics (31,32) However, the research participant have good impression on their personal experience with CE interventions, which align with study conducted by *Eva Turk and others*, acknowledge the advantages of CE interventions as an efficient method for responding to community needs (33) and as more sustainable strategy for dengue prevention and control (34–36). Nevertheless, our participants highlighted challenges such as the infodemic, lack of trust and financial constraints. These complexity resonate with finding from other researchers as barriers to CE interventions in conflict countries (37–39). Moreover, in Yemen door-to-door education strategy for vector-borne diseases have been used (40) evidence suggests a lack of community trust in the health system. participants also faced difficulties during door-to-door education in Aden governorate as result of spreading infodemic and security issues (39,41,42) Additionally, the cost of recruiting and training community health workers (37) raises the questions about the effectiveness of door-to-door education strategies for CE interventions in the current situation or for the conflict and post conflict context in general, and whether their effectiveness is differs between urban and rural areas.

### Outer setting domain

The conflict context presents numerous critical incidents that hinder health interventions(43). Security problems due to presence of different armed groups in Aden governorate (38) potential acute conflict causing transportation problems (37) and unstable political condition affecting health system policies and priorities are significant obstacles (30,44) Furthermore, critical incidence have led to increases vector-borne diseases, such as upsurge numbers of IDPs (45–48) and reported water and sanitation issues in Aden governorate (6,49–51). Lastly, the rapid spread of infodemic information has decreased health interventions effectiveness (41). For the local attitude construct, our literature search revealed no existing information to interoperate our results, either from Yemen context or other similar context except the population’s attitude toward utilizing traditional medicine (49,52).

The local condition of low economic and increase poverty (53) have negatively affected the community interest to health interventions (52). Qualitative case study in South Sudan and Haiti have highlighted negative behaviour changes in communities, such as losing community cohesion, lack of collaboration with community health workers and decreased desire to self-dependence due to inappropriate intervention design by international organization aid (54) This finding resonate with our research. Moreover, the weakness and politicization of Yemen’s health system (7,55) are consequences of political impacts. Additionally, the environmental condition, including rainfall are directly effecting CE intervention(37,39).

Partnerships and connections between government sectors and international organizations have been greatly affected by conflict(30).Health system fragmentation(7) and weakened trust in government sectors(32,54) have led to the current partnership style through health clusters organized by international organization(56).Nevertheless, the effectiveness of these health clusters is questionable (44). On the other hand, using social media tools such as

WhatsApp for connections shown success with similar context (57). The Lack of human resources, corruption, uncertainty among stakeholders(58) and the absence of health policies and organizational structure within Yemen’s health system (6,39) are major challenges. Before the conflict total health expenditure was low, with health spending per capita grew from 25$ in 2000 to 80$ in 2014,then falling to 75$ in 2015 (6). Currently, financing health program relies on donors supports (6,7) Political support and clear empowerment plan for the ministry of health with different donors and community support, are crucial to overcome financing problems (59,60).

### Inner setting domain

Governance fragmentation between Houthi in Northern Yemen and legitimate authority with the secessionist Southern Transitional Council (STC) in the south has weakened the health system (1,2) and led to lack of communication, management, and institutional work (6,7,61). Additionally, stress, increase workloads, and economical pressure are affecting the overall working culture (38,62). In conflict context, CE interventions are usually considered as low priority (63). Offering high incentives for community volunteers or exceeding the salary limits for health workers has negative long-term outcomes for the health system (37,64,65) similarly, to over-dependence on external aid (66). While, much evidence focuses on the importance of capacity building in the conflict context (30,37,57,67), national evidence shows clear gap (39,44). No evidence was available for relational connection or tension to change constructs.

### Individual domain

There is limited literature to discuss our research results related to the individual domain. Available evidence explains the implementers and opinion leaders view but not policymakers, However, our research participants indicate that health planning and donor fund allocation is mostly handled by the policymakers. National evidence highlights the role of governorate and district health office in implementing health interventions, with local authorities and governors as influencing factors (6). International evidence are emphasize the importance of involving traditional and community leaders(14,66). However, lack of motivation from community volunteers due to the economic needs is noted (37,68).

### Implementation processes domain

The implementation process is directly and indirectly affected by other four domains. Our findings suggests that coordination weaknesses between different level of authorities responsible for CE are mainly related to the health system politicization and the political unrest, as explained in the outer and inner domains (7,55,69,70). Research participants identified the need assessments to improve CE interventions (innovation delivery) due to lack of planning and capacity building, which resonates with other research findings as barriers in conflict settings (37,58,71). Participants perception of implementing CE interventions for dengue prevention and control are lack of recourse, and looking for personal benefit, these are considered as common consequences in conflict and post conflict context (37,58,72,73). Conversely, increased interest of CE and evidence of it is effectiveness in humanitarian contexts were also seen (74–76). Long-term planning or having a clear vison is challenging during fragile contexts (5,77)

The strategies suggested from by research participants to enhance CE interventions in dengue prevention and control such as political commitments to assure resources mobilization, aligned with research finding from Somalia and Afghanistan (78,79) and developing communication and coordination strategy which also mentioned in various international evidence (37,58,80) For better implementation and adoption of CE interventions, research participants suggested necessary modification to the current context (inner setting adaptation) Firstly, creating clear health policy for CE and SBC concepts to facilitate CE intervention work (74–76). Secondly, encourage community ownership and enhancing community role in health system to understand their needs and sustained outcome(31,80,80). The adaptation of CE interventions (innovation adaptation) suggestions includes the proper utilization of available human and material resources such as school students and religious people and political parties (14,34,35,40,81).Additionally, improving community communication massage by using more innovative and social media methods (71,82,83).

## Conclusions

The CE and SBC interventions are smart, affordable and resilience solution for many health problems in conflict-effected countries and it is essential to determine the best strategy according on available resources and to consider sustainability from early stages. Aden Governorate is impacted by the overall political situation, However, the cultural and religious value that emphasize the importance of helping others aligns with CE and social behavioural principles. Major barrier includes health system fragmentation, weak institutional work, low economic situations and a heavily reliance on funding agencies, facilitating factors include moral incentives, personal recognition, community charity by business owners, and providing the necessary trainings, which could improv the inner and outer settings. In conflict setting Individuals such as policymakers and influencers are playing dominant role. This dominance stems from overall systemic weaknesses which is affecting the implementation process of CE interventions throughout interference in participant selection and weak coordination with difficulty of having clear vison.

## Recommendation

1. Initiate community engagement and social behavioral change (SBC) interventions early
2. Involve community members in intervention design and planning to enhance community acceptance and ensure long-term sustainability.
3. Security issues should not hinder interventions
4. Interventions should promote cohesion between local communities and internally displaced persons (IDPs).
5. Strengthen existing laws and policies to improve collaboration between government sectors, agencies, and community leaders.
6. Ensure clear agreements between governments and donors to establish sustainable health plans, facilitating a smooth transition from relief to development phases.
7. Enhance institutional capacity by providing essential training and guidelines for community and SBC initiatives.
8. Adaptation is an essential component for the functionality, accountability, and data-driven improvement of health programs.

### Research strengths & limitations

To the best of the researcher knowledge, this the first implementation research in Yemen using CFIR framework. The research investigator’s experience as a malaria program coordinator for two years was a significant facilitator in developing the research proposal and data collection process, and served ad continues motivator throughout the study.

However, the research faced some limitation in both data collection and data analysis. During data collection, we were unable to meet with the responsible team of Social and Behavioural Change (SBC) program in Aden UNICEF office, despite continuous formal and informal request. UNICEF is considered as a leading organization of CE and SBC in Yemen and was the first to suggested using SBC concept in humanitarian context globally (74). The data analysis conducted solely by research investigator, with guidance from the internal and external advisors. Time constraints and resources limitation was a significant challenge.

### Future research directions

We think there is a research gap of understanding the perception of the key stakeholders regarding the exact meaning of community engagement and social behavioural change approach. Additionally, further research is needed to explore the degree of political influence on health system in the conflict and post-conflict contexts.

## Data Availability

All data produced in the present study are available upon reasonable request to the authors

## Authors’ contributions

MMD (Mustafa Mohammed Dhaiban) conceptualized the study, conducted the research, performed data collection and analysis, and prepared the initial manuscript draft. HB (Dr. Huda Basaleem) provided guidance during the development of the research proposal and contributed to formulating the initial draft of the manuscript. NP (Dr. Neetu Purohit) assisted with revising the final draft of the manuscript, enhancing the analysis of results, and providing critical input on the interpretation of findings. HB and NP served as key guides throughout the research process. Additionally, HB (Dr. Huda Basaleem) and NP (Dr. Neetu Purohit) acted as external and internal advisors, respectively, offering oversight and valuable guidance as supervisors of MD’s master’s research, which formed the basis of this manuscript.

## Acknowledgements

The authors extend their gratitude to the TDR program for providing the scholarship that supported this study and to IIHMR University’s S.D. Gupta School of Public Health for its academic and logistical assistance throughout the research process.

